# COVID-19 vaccines effectiveness against SARS-CO-V-2 infection among persons attending RT-PCR centre at a Medical College Hospital in Telangana: A case control study

**DOI:** 10.1101/2022.05.15.22273945

**Authors:** Nagapraveen Veerapu, Dhanashree P Inmdar, Baer P Ravi Kumar, Basavaraju Anuradha, Pavitra Guddanti, Sree D Issapuri, Nikhita S Ganta, Arun Gopi

## Abstract

**Background:** In January 2021, India drug regulator issued restricted emergency approval for COVAXIN and COVISHIELD which were manufactured in India. On mid-January 2021, in India, there were 10.5million confirmed cases and 0.15 million deaths.

**Objectives:** The objectives were to evaluate vaccine effectiveness (VE) of India made Covid-19 vaccines against SARS-CoV-2 infection.

**Methods:** A test negative case control study was conducted from May 2021 to December 2021 for duration of 8months among people attending an RT-PCR centre at a medical college Hospital for RT-PCR test. The baseline characteristics and RT-PCR report; and preliminary data about vaccine status were collected from the RT-PCR centre. The exposure to vaccination was enquired via Phone call or was checked with data available with the health authorities.

**Results:** After applying inclusion exclusion criteria, case and control definitions, a total of 380 participants (95cases and 285 controls) were included. The adjusted VE of two doses of COVISHIED vaccine against symptomatic SARS-CoV-2 infection was 52.2% (95% CI, 41.7 to 62.1) and single dose was 40.88% (95% CI, 31.26 to 51.29). The adjusted VE of two doses of COVAXIN vaccine against SARS-CoV-2 infection was 39% (95% CI, 29.40 to 49.27). The overall VE was 48.20% (95% CI, 37.90 to 58.22) for two doses of any vaccines.

**Conclusions:** India made vaccines were nearly 50% effective. Similar results show by different studies with a margin of 10-25% difference. Further new studies should be conducted as new variants of SARS-CoV-2 are emerging, and we don’t know how the vaccine works against the variants and booster doses were required or not.

## Introduction

Coronavirus disease (COVID-19) is an infectious disease caused by a newly discovered coronavirus.^1^ The Causative agent is SARS-CoV-2.^1,2^ The outbreak was declared as a Public Health Emergency of International concern on 30 January 2020.^3^ World Health Organization (WHO) on 11th March 2020 declared Covid-19 disease a pandemic.^4^

On Mid Jan 2021, as per WHO situation update report on Novel Coronavirus Disease-worldwide there were 93.2 million confirmed cases and 2million deaths. In South East Asian Region there were 12.5 million confirmed cases and 1.9 lack deaths. In India, there were 10.5million confirmed cases and 1.5lack deaths.^5^

In January 2021, India’s drug regulator issued restricted emergency approval for COVAXIN and COVISHIELD which were manufactured in India. COVISHIELD vaccine is a Recombinant Chimpanzee Adenovirus vector vaccine.^6^ Its efficacy against symptomatic SARS-CoV-2 infection after two doses was 76%.^7^ COVAXIN is a Whole Virion Inactivated Corona Virus Vaccine developed in collaboration with ICMR.6 The Bharat Biotech BBV152 COVAXIN vaccine efficacy against COVID-19 after two doses was 78%.^8^ On 16 January 2021, India launched the largest vaccine drive against COVID-19 disease.^9^ Vaccine effectiveness (VE) is a measure of how well vaccines works in the real-world settings. On 23 March 2021, the interim WHO guidance how to evaluate COVID-19 vaccine effectiveness (VE) was provided based on previous guidance on VE evaluations.^10^ The objectives were to evaluate effectiveness of different India made Covid-19 vaccines (COVISHIELD & COVAXIN) and to evaluate overall effectiveness in India made COVID-19 vaccines against symptomatic SARS-CoV-2 infection.

## Material and methods

A test negative case control study was conducted from May 2021 to December 2021, 8months duration among people attending an RT-PCR centre at a medical college Hospital in Khammam, Telangana for Reverse transcriptase Polymerase chain reaction (RT-PCR) test. The persons who were 18 years and above age and who gave informed oral consent will be included in the study. The persons who were not able co-operate, not willing to participate, pregnant women, administered with other Covid vaccines were excluded from the study.

A Case was a person who had at least one Covid-19 symptoms and laboratory confirmed-tested positive for SARS-CoV-2 infection for RT-PCR test. The range of symptoms are fever, cough, shortness of breath, fatigue, body aches, headache, new loss of taste or smell, sore throat, runny nose, vomiting and diarrhoea.^11^ A control was defined as a person who had no symptoms of Covid-19 disease and seeking the test for other purposes like admission to medical & surgical wards, college admissions and travel; and also tested negative for SARS-CoV-2 infection for RT-PCR test. In a week, for every 4-5 cases and 12 to 15 controls were chosen and age group matching was done for data collection period (24 weeks). Cases to controls ratio were 1:3.

The Baseline characteristics and RT-PCR laboratory report; and preliminary data about vaccine status were collected from the RT-PCR centre. The exposure to vaccination was noted from the data available with the district health authorities or enquired via phone call. The data given by the persons during the interview was considered if the data was not found with the health authorities or by mobile phone call. Data-collection forms were assessable only to the principal investigators and confidentiality was maintained.

Vaccine status was taken on the day of RT-PCR testing. A person was considered as vaccinated with first dose-after 14days of vaccination; and a person was considered as vaccinated with second dose-after 7days of vaccination. A person who had received only first dose was considered as partially vaccinated and a person who had received second dose (2 doses) was considered as completely vaccinated person. Ethical clearance was obtained from Institutional Ethics committee.

### Statistical Analysis

First the data is entered in Microsoft Excel and then transformed into R software version 4.1.2 and analysed. The data was represented by frequency, percentage, mean and standard deviation. Conditional logistic regression was utilized to find vaccine effectiveness, 1 minus odds ratio (×100) for complete and partial vaccination compared against no vaccination in India made vaccines. The vaccine effectiveness was calculated at 95% confidence interval. The final model was adjusted for age and gender. Adjusted vaccine effectiveness had been calculated based on adjusted odds ratios in the final model.

## Results

After applying the inclusion and exclusion criteria, case and control definitions, a total of 380 participants (95cases and 285 controls) were included. Out of 380 participants, 194 (51.1) were males and 186 (48.9%) were females. All participants’ age was in the range of 18years to 92years with the mean age of 41.98 ± 16.88.

The unadjusted VE of single dose and two doses of COVISHIED vaccine against symptomatic SARS-CoV-2 infection was 40.88% (95% CI, 31.26 to 51.29) and 62.64% (95% CI, 52.76 to 72.44); and the adjusted VE for single dose and two doses was 31.1% (95% CI, 22.10 to 41.03) and 52.2% (95% CI, 41.7 to 62.1) respectively. The unadjusted and adjusted VE for two doses of COVAXIN was 50% (95% CI, 39.83 to 60.17) and 39% (95% CI, 29.40 to 49.27). The overall unadjusted and adjusted VE in India made vaccines was 58.81% (95% CI, 48.70 to 68.70) and 48.20% (95% CI, 37.90 to 58.22) respectively. The completely vaccinated participants VE was more than partial vaccination as compared to unvaccinated participants. (Table 1).

**Table 1:**
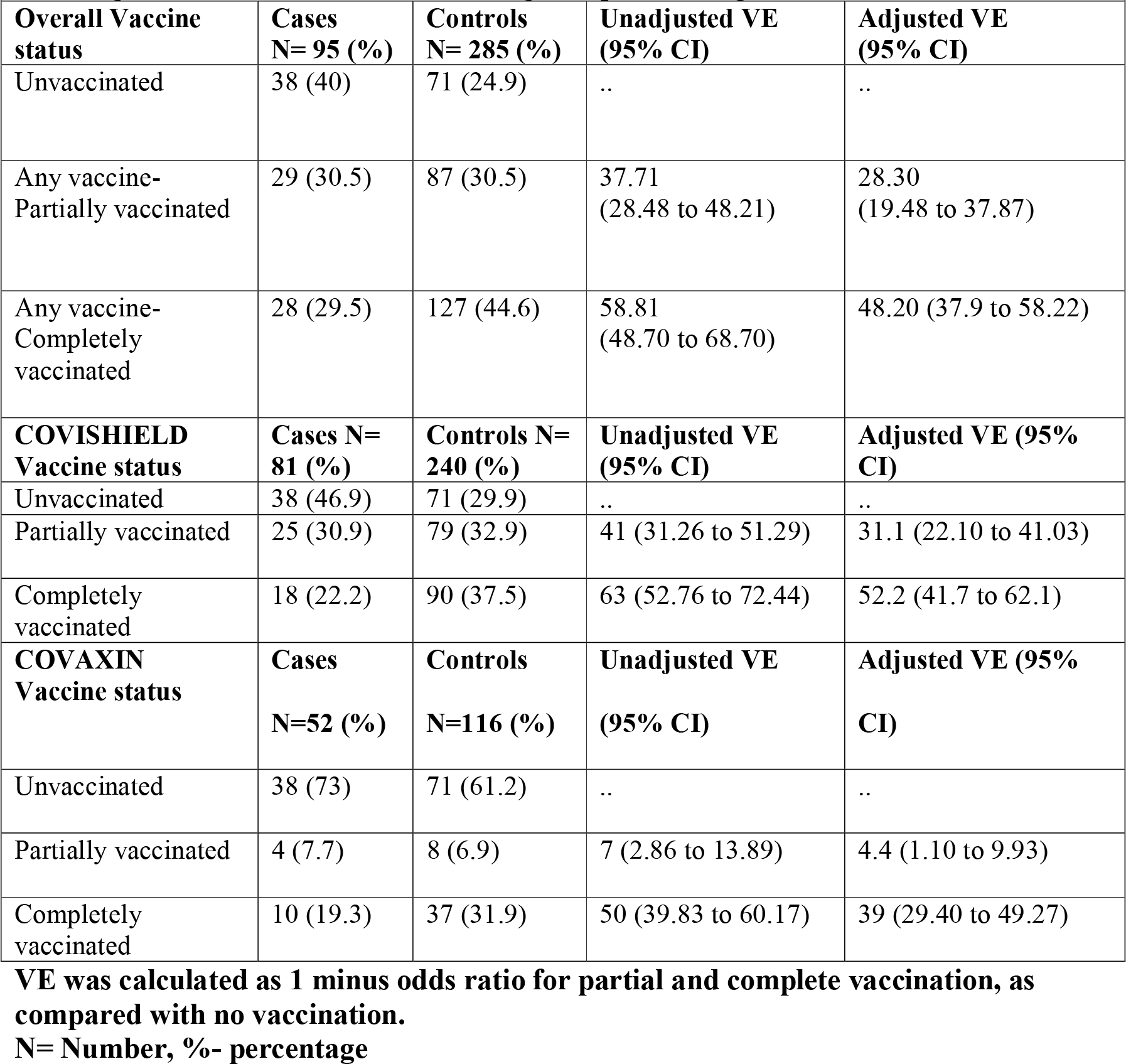
Evaluation of Overall VE, COVISHIELD and COVAXIN VE among the participants attending RT-PCR centre at a medical college Hospital, Telangana.

| **attending RT-PCR centre at a medical college Hospital, Telangana** |  |  |  |  |
| --- | --- | --- | --- | --- |
| **Overall Vaccine status** | **Cases N= 95 (%)** | **Controls N= 285 (%)** | **Unadjusted VE (95% CI)** | **Adjusted VE (95% CI)** |
| Unvaccinated | 38 (40) | 71 (24.9) | .. | .. |
| Any vaccine-Partially vaccinated | 29 (30.5) | 87 (30.5) | 37.71<br>(28.48 to 48.21) | 28.30<br>(19.48 to 37.87) |
| Any vaccine-Completely vaccinated | 28 (29.5) | 127 (44.6) | 58.81<br>(48.70 to 68.70) | 48.20 (37.9 to 58.22) |
| **COVISHIELD Vaccine status** | **Cases N= 81 (%)** | **Controls N= 240 (%)** | **Unadjusted VE (95% CI)** | **Adjusted VE (95% CI)** |
| Unvaccinated | 38 (46.9) | 71 (29.9) | .. | .. |
| Partially vaccinated | 25 (30.9) | 79 (32.9) | 41 (31.26 to 51.29) | 31.1 (22.10 to 41.03) |
| Completely vaccinated | 18 (22.2) | 90 (37.5) | 63 (52.76 to 72.44) | 52.2 (41.7 to 62.1) |
| **COVAXIN Vaccine status** | **Cases N=52 (%)** | **Controls N=116 (%)** | **Unadjusted VE (95% CI)** | **Adjusted VE (95% CI)** |
| Unvaccinated | 38 (73) | 71 (61.2) | .. | .. |
| Partially vaccinated | 4 (7.7) | 8 (6.9) | 7 (2.86 to 13.89) | 4.4 (1.10 to 9.93) |
| Completely vaccinated | 10 (19.3) | 37 (31.9) | 50 (39.83 to 60.17) | 39 (29.40 to 49.27) |

**VE was calculated as 1 minus odds ratio for partial and complete vaccination, as compared with no vaccination.**

The overall adjusted VE of the participants for two doses in the age group 41-64years was 68.8% (95% CI, 58.97 to 77.87) and was more than the other age groups, 18-40years was 45.5% (95% CI, 35.03 to 55.27) and ≥65 years was 13.4% (95% CI, 7.11 to 21.2). The COVISHIELD VE of the participants for 2 doses in the age group≥65 years was 69.1% (95% CI, 58.92 to 77.87) and was more than the other groups, 41-64years was 58.3% (95% CI, 47.72 to 67.8) and 18-40years was 43.2% (95% CI, 33.1 to 53.2) (Table 2 & 3). The overall adjusted VE of the participants for two doses for males was 55.2% (95% CI, 44.73 to 64.97) and was more than females (95% CI, 39.1% (29.4 to 49.27)). The COVISHIELD VE of the participants for 2 doses for males was 55.7 (95% CI, 45.72 to 65.92) and was more than females and was 48.3% (95% CI, 37.90 to 58.22) (Table 2 & 3). COVAXIN VE according to age and gender was not quantified due to small sample size.

**Table 2:** Evaluation of overall VE among participants attending RT-PCR centre according to age and gender.

**VE was calculated as 1 minus odds ratio for partial and complete vaccination, as compared with no vaccination.** **N= Number, %- percentage**
|  |  |  |  |  |
| --- | --- | --- | --- | --- |
| **Matched Age groups (years)** | **Cases N=95 (%)** | **Controls N= 285 (%)** | **Unadjusted VE (95% CI)** | **Adjusted VE (95% CI)** |
| **18-40** |  |  |  |  |
| Unvaccinated | 19 (20) | 35 (12.3) | .. | .. |
| Partially vaccinated | 15 (15.8) | 50 (17.5) | 44.8 (34.08 to 54.28) | 34.5 (24.82 to 44.15) |
| Completely vaccinated | 14 (14.7) | 59 (20.7) | 56.3 (45.72 to 65.92) | 45.5 (35.03 to 55.27) |
| **41-64** |  |  |  |  |
| Unvaccinated | 16 (16.9) | 26 (9.1) | .. | .. |
| Partially vaccinated | 10 (10.5) | 30 (10.5) | 45.9 (35.98 to 56.26) | 34.4 (24.82 to 44.15) |
| Completely vaccinated | 10 (10.5) | 52 (18.3) | 68.8 (58.97 to 77.87) | 57.7 (47.71 to 67.8) |
| **≥ 65** |  |  |  |  |
| Unvaccinated | 3 (3.2) | 10 (3.5) | .. | .. |
| Partially vaccinated | 4 (4.2) | 7 (2.5) | NE | NE |
| Completely vaccinated | 4 (4.2) | 16 (5.6) | 16.7 (10.23 to 25.82) | 13.4 (7.11 to 21.2) |
| **Gender** | **Cases N= 95 (%)** | **Controls N= 285 (%)** | **Unadjusted VE (95% CI)** | **Adjusted VE (95% CI)** |
| **Males** |  |  |  |  |
| Unvaccinated | 21 (22.1) | 33 (11.6) | .. | .. |
| Partially vaccinated | 10 (10.5) | 44 (15.4) | 64.3 (53.79 to 73.36) | 52.4 (41.78 to 62.1) |
| Completely vaccinated | 15 (15.8) | 71 (24.9) | 66.8 (55.85 to 75.18) | 55.2 (44.73 to 64.97) |
| **Females** |  |  |  |  |
| Unvaccinated | 17 (17.9) | 38 (13.3) | .. | .. |
| Partially vaccinated | 19 (20) | 43 (15.1) | 1.3 (0.02 to 5.4) | 0.9 (0.40 to 1.64) |
| Completely vaccinated | 13 (13.7) | 56 (19.7) | 48.2 (37.9 to 58.22) | 39.1 (29.4 to 49.27) |

**VE was calculated as 1 minus odds ratio for partial and complete vaccination, as compared with no vaccination.**

**Table 3:** Evaluation of COVISHIELD VE among participants attending RT-PCR centre according to age and gender.

| **Matched Age groups (years)** | **Cases N= 81 (%)** | **Controls N= 240 (%)** | **Unadjusted VE (95% CI)** | **Adjusted VE (95% CI)** |
| --- | --- | --- | --- | --- |
| **18-40** |  |  |  |  |
| Unvaccinated | 19 (23.5) | 35 (14.6) | .. | .. |
| Partially vaccinated | 13 (16) | 45 (18.8) | 46.8 (36.92 to 57.24) | 36.3 (26.6 to 46.21) |
| Completely vaccinated | 10 (12.5) | 40 (16.7) | 54 (43.72 to 64.0) | 43.2 (33.1 to 53.2) |
| **41-64** |  |  |  |  |
| Unvaccinated | 16 (19.8) | 26 (10.8) | .. | .. |
| Partially vaccinated | 9 (11) | 27 (11.3) | 45.9 (35.98 to 56.26) | 34.4 (24.82 to 44.15) |
| Completely vaccinated | 7 (8.6) | 37 (15.4) | 69.3 (58.92 to 77.8) | 58.3 (47.72 to 67.8) |
| **≥ 65** |  |  |  |  |
| Unvaccinated | 3 (3.7) | 10 (4.2) | .. | .. |
| Partially vaccinated | 3 (3.7) | 7 (2.9) | NE | NE |
| Completely vaccinated | 1 (1.2) | 13 (5.3) | 74.4 (64.27 to 82.26) | 69.1 (58.92 to 77.87) |
| **Gender** | **Cases**<br>**N=81** | **Controls**<br>**N=240** | **Unadjusted VE (95%**<br>**CI)** | **Adjusted VE (95%**<br>**CI)** |
| **Males** |  |  |  |  |
| Unvaccinated | 21 (25.9) | 33 (13.8) | .. | .. |
| Partially vaccinated | 9 (11.1) | 39 (16.3) | 63.8 (53.72 to 73.36) | 51.8 (41.78 to 62.10) |
| Completely vaccinated | 10 (12.3) | 48 (20) | 67.3 (56.88 to 76.08) | 55.7 (45.72 to 65.92) |
| **Females** |  |  |  |  |
| Unvaccinated | 17 (21) | 38 (15.8) | .. | .. |
| Partially vaccinated | 16 (19.8) | 40 (16.6) | 10.6 (5.62 to 18.83) | 7.6 (3.52 to 15.16) |
| Completely vaccinated | 8 (9.9) | 42 (17.5) | 57.5 (46.71 to 66.86) | 48.3 (37.90 to 58.22) |

## Discussion and conclusions

The VE of India made vaccines was nearly 50%. The VE (two doses) of COVISHIELD (52.2%) was more than COVAXIN (39%) by 13.2%. Similar results were show by different studies conducted in India with a margin of 10-25% difference in the VE.

VE in our study for partial and complete vaccination of COVISHIELD was 31.1% (95% CI, 22.10 to 41.03) and 52.2% (95% CI, 41.7 to 62.1). In a study conducted in Faridabad showed VE of two doses of ChAdOx1 nCoV-19 (COVISHIELD) against SARS-CoV-2 infection was more (63·1%) and single dose against SARS-CoV-2 infection was more (46·2%).^12^ In a study from CMC Vellore, VE for single dose and two doses was 61% and 65% respectively and it was more than our study.^13^ In a study done at Gangaram Hospital, New Delhi during surge of infection, the VE after single dose and two doses of COVISHIELD was 18% and 28% respectively and it was less than our study, might be due to peaking of the pandemic.^14^ The studies outside India showed VE similar to our study. In Sao Paul, VE for single dose of ChAdOx1 nCoV-19 vaccination was 33.4% after 28 days of administration.^15^ In a Scottish study, VE was 60% for delta variants after two doses, and was more than our study.^16^ In study conducted in AIIMS Delhi the effectiveness of two doses COVAXIN BBV152 against symptomatic COVID-19 was 46% after 28days of vaccination, in our study VE was 39% but it was after 14days of vaccination. In our study, VE of single dose of COVAXIN was not much effective (6.1%) and was similar to AIIMS study. The overall VE among males is more than females but, in a study, conducted in AIIMS the VE for COVAXIN among females was (66%) more than males (38%).^17^

The VE of m-RNA vaccines in was higher as compared to made in India vaccines (COVAXIN & COVISHIELD). The Overall VE of mRNA vaccines (BNT162b2 vaccine and the mRNA-1273) was 90.4%.^18^

A prospective study in Scotland the overall VE after first dose of BNT162b2 mRNA vaccine and ChAdOx1 nCoV-19 was estimated to prevent hospital admissions due to Covid-19 disease was more than 60%.^19^ but our study estimated the VE against symptomatic SARS-Co-V-2 infection, which is nearly 50% for 2 doses.

There were several limitations like sample size was small and some factors like covid appropriate behaviour, socio-economic status, occupation were not considered. Further, more studies should be conducted as new variants of SARS-CoV-2 like Omicron are emerging, and we don’t know how the vaccine works against the variants and whether booster doses are required or not.

## Data Availability

All data produced in the present study available upon reasonable request to the authors.

## Acknowledgement

Nil

